# Reliability of retrospective self-report of shift work exposure in a large occupational cohort

**DOI:** 10.64898/2026.10.27.26366313

**Authors:** Clara López-Velasco

## Abstract

**Objectives:** To examine agreement between baseline and retrospective assessments of shift work exposure and test whether this differs in cancer and cardiometabolic disease patients.

**Methods:** A subgroup of the Airwave Health Monitoring study, a cohort of police employees, self-reported shift work status at study entry and retrospectively at follow-up. Agreement statistics were computed overall and stratified by subsequent cancer and cardiometabolic disease status. Adjusted logistic regression tested whether discordant reporting differed by disease status.

**Results:** Overall agreement was 86% (95% CI = 84–88%; κ = 0.72), with no evidence of asymmetric misclassification (McNemar’s p = 0.32). Agreement was similar among participants who later developed cancer (κ = 0.76) but lower, although imprecisely estimated, among those who developed cardiometabolic disease (κ = 0.56). Adjusted odds of discordant reporting did not differ significantly by cancer (OR = 0.96, 95% CI = 0.39–2.05) or cardiometabolic disease status (OR = 1.85, 95% CI = 0.81–3.87).

**Conclusions:** Retrospective shift work reports showed good overall reliability, with no evidence of asymmetric misclassification, though agreement point estimates varied somewhat by disease outcome. These findings provide an empirical basis for future studies using retrospective shift work exposure in the Airwave Health Monitoring Study and similar cohorts.

## Introduction

Shift work has been associated with a range of adverse health outcomes, from cardiometabolic diseases to cancers, although evidence for these associations remains inconsistent (1,2). A challenge in research on shift work and health is the reliable ascertainment of shift work exposure. As research works to establish whether shift work causes ill health (and if so, for which conditions, in whom, and through which pathways and schedules) this rests on how accurately shift work exposure itself is measured.

Large population cohorts typically rely on self-report, and some introduce a retrospective element by asking, at follow-up, whether participants did shift work at an earlier point in time. This is the case for the Airwave Health Monitoring Study (AHMS), a large occupational cohort of police officers and staff (3). However, an issue in the use of retrospective reports is that the recall of occupational exposures can degrade over time, especially for complex shift work systems (4,5).

The accuracy of retrospective reports might also depend on a participant’s subsequent health status, a phenomenon known as differential recall bias (6). This bias has long been established in epidemiology but is only beginning to be examined in shift work research. For example, Vestergaard et al. (7) found that breast cancer patients had a better recall of ever having done night shift work compared to controls, and correcting for this misclassification produced more conservative shift work exposure risk estimates.

The aims of this report are twofold: to quantify agreement between baseline and retrospective shift-work reporting in the AHMS, and to test whether agreement differs by eventual disease status, separately for cancer and cardiometabolic disease. This report aims to provide empirical evidence to inform whether, and under what conditions, retrospectively reported shift work exposure can reasonably be treated as a proxy for baseline exposure in future AHMS analyses.

## Methods

This report focuses on a subset of participants (n = 1,863) from the AHMS (3) who completed a shift work exposure assessment both at study entry (baseline) and retrospectively at follow-up (see participant flowchart, Figure S1). In the baseline questionnaire, they were asked whether they were a shift worker, being defined as working “outside the regular daytime hours of approximately 7am and 6pm”. At follow-up, they were asked whether they were a shift worker in the year of entry, providing the same definition of shift work. Those reporting working shifts only two or three times a year were coded as day workers. A sensitivity analysis additionally examined agreement using the original three-category variable (no shift work, two or three times a year, regular shift work).

Cardiometabolic disease cases were defined and identified through linkage with national hospital admissions data, based on ICD-10 codes for stroke (I60–I64), coronary artery disease (I20–I25), and heart failure (I50) as primary admissions, and diabetes (E10–E14) as primary or secondary admissions. Cancer cases were identified through linkage with national cancer registry data, including malignant cancers at all sites except non-melanoma skin cancer (C00–C97, excluding C44). To maximize power, primary analyses included all incident cases arising between study entry and end of linked data availability (31 October 2024 for cardiometabolic disease, n = 41; 31 December 2023 for cancer, n = 58). A sensitivity analysis restricted cases to those diagnosed prior to follow-up completion (n = 14 for cardiometabolic disease and n = 17 for cancer).

We computed the agreement, sensitivity and specificity of retrospectively reported shift work status compared to that reported at baseline, along with 95% confidence intervals, Cohen’s κ coefficients (to test agreement adjusted for chance), and McNemar’s p-value (to test for directionality of discordant reporting). McNemar’s test was substituted by Bowker’s test of symmetry for three-category analysis. To examine whether recall agreement depends on subsequent health status, the same agreement statistics were computed stratified by cancer and cardiometabolic disease status. To predict discordant reporting across disease status, a logistic regression model was used for each outcome, adjusted for age at follow-up (continuous), sex (female, male), ethnicity (white, other), occupational rank at entry (police staff, police officer, other), and years from entry to follow-up (continuous). Analyses were completed in R, version 4.5.3.

## Results

Sociodemographic characteristics of the study cohort are described in Table S1. The average number of years between baseline and follow-up was 4.10 (SD = 0.97). The agreement between retrospective and baseline shift work status reports was 86% (84%– 88%), as presented in Table 1 (see Table S2 for a cross-classification table). Using baseline data as the reference, the sensitivity of retrospective shift work data was 88% (86%–90%; the proportion of baseline shift workers that were identified at follow-up), while the specificity was 84% (81%–86%; the proportion of baseline non-shift workers identified). A Cohen’s κ of 0.72 (0.69–0.75) suggests a substantial agreement between timepoints (8). A non-significant McNemar’s test (p = 0.32) indicates no statistically detectable asymmetry in discordant switching between “yes” and “no” responses. A sensitivity analysis using the original three-category variable (day work only, two or three times a year, regular shift work) showed markedly lower agreement for the two or three times a year category (Table S3), supporting the decision to collapse this category into non-shift-work status in the main analysis.

**Table 1.**
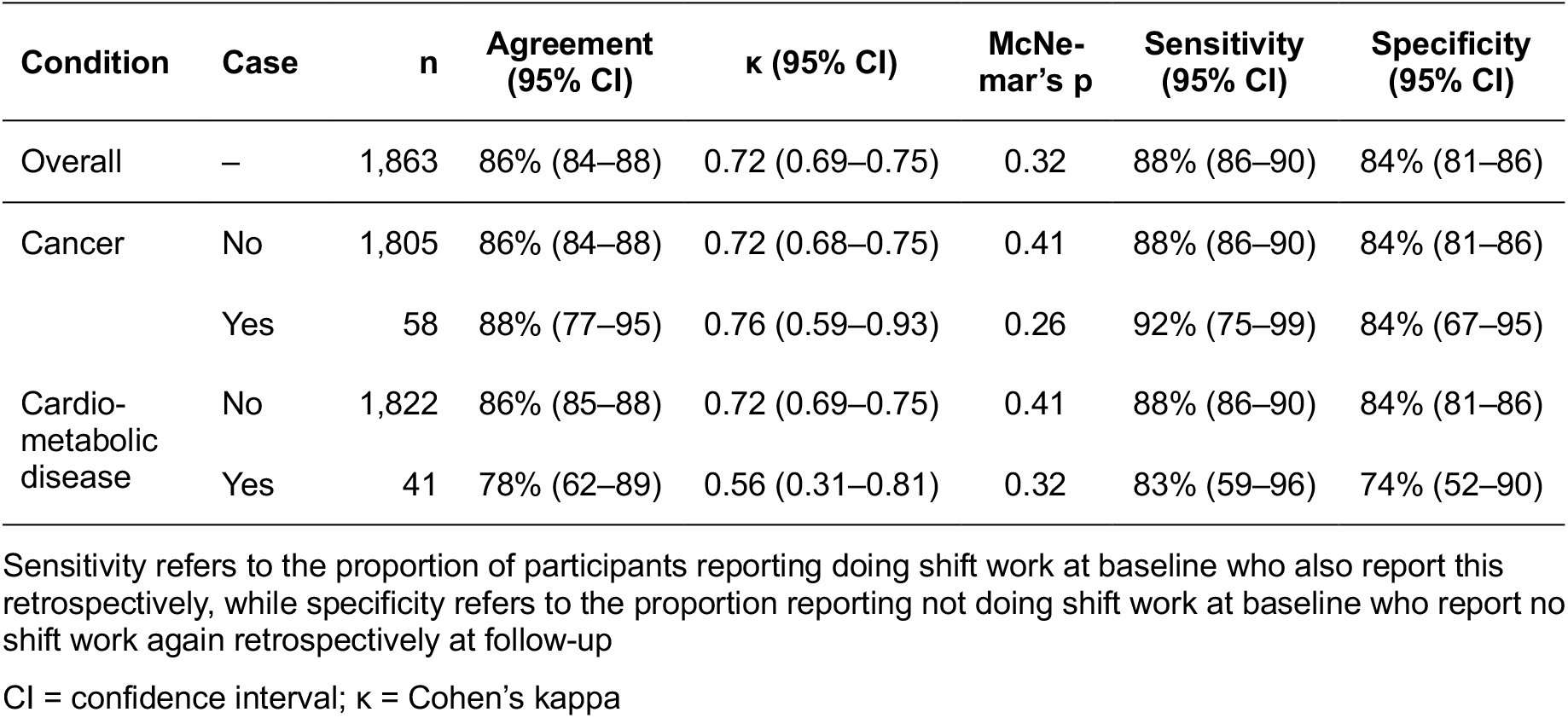
Agreement between baseline and retrospectively reported shift work status, overall and by subsequent cancer and cardiometabolic disease status.

Raw agreement statistics were broadly similar for cancer patients but appeared lower for cardiometabolic disease patients, although confidence intervals were wide and overlapped with those for non-cases (Table 1). McNemar’s results remained non-significant across all subgroups, indicating no evidence of asymmetric misclassification in either cancer or cardiometabolic disease patients. Further, after adjustment for age, sex, ethnicity, rank, and years between baseline and follow-up, the odds of discordant reporting did not differ for cancer (OR = 0.96, 95% CI = 0.39–2.05) or cardiometabolic disease patients (OR = 1.85, 95% CI = 0.81–3.87). In the sensitivity analysis restricted to cases diagnosed before follow-up, agreement was 88% for cancer (n = 17) and 71% for cardiometabolic disease (n = 14), with wide confidence intervals (Table S4).

## Discussion

This report examined the agreement between baseline and retrospective assessments of shift work exposure, and whether this differs in cancer and cardiometabolic disease patients within the AHMS, an occupational cohort of police officers and staff.

Overall agreement between baseline and retrospective shift work reports was good (86%; κ = 0.72), and consistent with similar assessments in the shift work literature (5). However, recall accuracy may not be uniform across shift work subtypes: Härmä et al. (4) found higher estimates for shift work involving night shifts (κ = 0.86) than for shift work without nights (κ = 0.42). Given the general shift work definition of the AHMS, our overall agreement might obscure similar heterogeneity.

Participants who subsequently developed cancer had agreement comparable to non-cases. Our results contrast with Vestergaard et al. (7), who found better recall of night shift work in breast cancer patients, although the studies are not directly comparable given different case and exposure definitions.

Point estimates of agreement, sensitivity and specificity were lower among participants who later developed cardiometabolic disease, but confidence intervals overlapped with those of non-cases and adjusted odds of discordant reporting did not differ significantly. These two forms of differential misclassification would be expected to act in opposite directions, with a lower sensitivity reducing shift work estimates, and a lower specificity inflating them (9,10). Given these opposing effects and the small sample size, future research using quantitative bias analysis in larger samples is needed to assess this.

These findings should be considered taking certain limitations into account. First, police employees are a unique occupational group with high shift work exposure (11) and tolerance and might only be representative of similar occupational cohorts. Second, retrospective recall data was compared with self-reported data collected at baseline and could not be validated against objective measures such as payroll data, the gold standard (4). Third, the average number of years between baseline and follow-up assessments was only four years, and the reliability of retrospective shift work reports could deteriorate in longer time periods. Despite this, the number of years between baseline and follow-up was not a significant predictor of discordant reporting in this study (data not shown).

To conclude, we found that retrospective self-reports of shift work showed good agreement with reports collected at study entry, with no evidence of asymmetric misclassification.

Agreement was similar for participants who later developed cancer, whereas point estimates were somewhat lower among those who developed cardiometabolic disease, although differences were not statistically significant. In both cases, there was no evidence of directional bias, suggesting that any misclassification present is unlikely to systematically distort shift work exposure risk estimates. Future research should confirm this using quantitative bias analysis once larger samples are available. We hope that future studies on shift work and health in the AHMS and similar cohorts can draw on these agreement statistics to consider whether the use of retrospectively reported shift work exposure is appropriate in their research context.

## Data Availability

The data used in this study were obtained from the Airwave Health Monitoring Study. The data are not publicly available but can be accessed by bona fide researchers upon request and approval from the Airwave Study Management Committee.

## Acknowledgements

Thanks to the Airwave Health Monitoring Study participants, police forces, and study team for their contribution. The Airwave Health Monitoring Study received ethics approval from the National Health Service Multi-Site Research Ethics Committee (MREC/13/NW/0588). All participants gave written informed consent before taking part. The author declares no conflicts of interest.

