## Supplemental Data 1 for "Reliability of retrospective self-report of shift work exposure in a large occupational cohort"

**Supplementary Materials** for *Reliability of retrospective self-report of shift work exposure in a large occupational cohort*

**Table 1**. Agreement between baseline and retrospectively reported shift work status, overall and by subsequent cancer and cardiometabolic disease status.

| Condition | Case | n | Agreement (95% CI) | κ (95% CI) | McNemar's p | Sensitivity (95% CI) | Specificity (95% CI) |
|---|---|---|---|---|---|---|---|
| Overall | – | 1,863 | 86% (84–88) | 0.72 (0.69–0.75) | 0.32 | 88% (86–90) | 84% (81–86) |
| Cancer | No | 1,805 | 86% (84–88) | 0.72 (0.68–0.75) | 0.41 | 88% (86–90) | 84% (81–86) |
| | Yes | 58 | 88% (77–95) | 0.76 (0.59–0.93) | 0.26 | 92% (75–99) | 84% (67–95) |
| Cardiometabolic disease | No | 1,822 | 86% (85–88) | 0.72 (0.69–0.75) | 0.41 | 88% (86–90) | 84% (81–86) |
| | Yes | 41 | 78% (62–89) | 0.56 (0.31–0.81) | 0.32 | 83% (59–96) | 74% (52–90) |

Sensitivity refers to the proportion of participants reporting doing shift work at baseline who also report this retrospectively, while specificity refers to the proportion reporting not doing shift work at baseline who report no shift work again retrospectively at follow-up

CI = confidence interval; κ = Cohen's kappa

**Figure S1**. Participant Flowchart

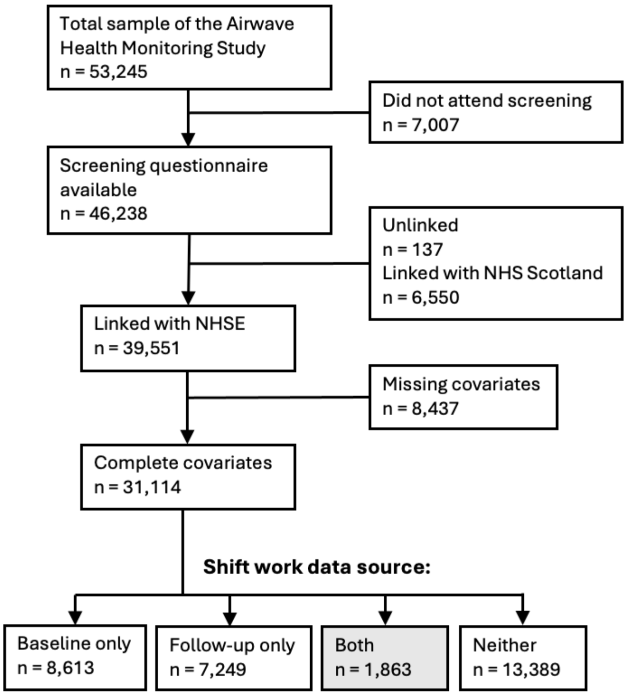


This report focuses on the n = 1,863 participants with shift work data available at both baseline and follow-up

**Table S1**. Characteristics of participants with shift work data at both timepoints compared with those with data at only one timepoint

| Characteristic | Source of shift work data | | SMD | 95% CI for SMD |
|---|---|---|---|---|
| | **One timepoint only (baseline or follow-up)**[a] N = 15,862 | **Both timepoints (analytic sample)** N = 1,863 | | |
| Age at Baseline | 40.8 (8.7) | 42.8 (8.1) | -0.24 | -0.28, -0.19 |
| Age at Follow-up | 48.9 (9.1) | 46.9 (8.1) | 0.24 | 0.19, 0.29 |
| Missing | 7,943 | 0 | | |
| Sex | | | 0.17 | 0.12, 0.22 |
| Female | 5,702 (36%) | 522 (28%) | | |
| Male | 10,160 (64%) | 1,341 (72%) | | |
| Ethnicity | | | 0.09 | 0.04, 0.14 |
| White | 14,697 (93%) | 1,680 (90%) | | |
| Other | 1,165 (7.3%) | 183 (9.8%) | | |
| Job Rank | | | 0.04 | 0.00, 0.09 |
| Police Staff | 4,133 (26%) | 473 (25%) | | |
| Police Officer | 11,477 (72%) | 1,369 (73%) | | |
| Other | 252 (1.6%) | 21 (1.1%) | | |
| Income | | | 0.48 | 0.43, 0.53 |
| £25,999 or less | 2,364 (15%) | 121 (6.5%) | | |
| £26,000–37,999 | 6,097 (38%) | 467 (25%) | | |
| £38,000–59,999 | 6,726 (42%) | 1,097 (59%) | | |
| £60,000 or more | 675 (4.3%) | 178 (9.6%) | | |
| Education | | | 0.07 | 0.02, 0.12 |
| L2 qualifications or below | 6,347 (40%) | 699 (38%) | | |
| L3 qualifications (secondary) | 4,893 (31%) | 560 (30%) | | |
| L4+ qualifications (higher) | 4,622 (29%) | 604 (32%) | | |
| Shift Worker (baseline) | 5,152 (60%) | 1,019 (55%) | 0.10 | 0.05, 0.15 |
| Missing | 7,249 | 0 | | |
| Shift Worker (follow-up) | 4,495 (62%) | 1,035 (56%) | 0.13 | 0.08, 0.18 |
| Missing | 8,613 | 0 | | |

[a] "One timepoint only" includes 8,613 participants with baseline shift work data only and 7,249 with follow-up data only.

Mean (SD) for continuous variables; n (%) for categorical variables. SMD = standardized mean difference, CI = confidence interval.

**Table S2.** Cross-classification of shift work status at baseline and follow-up.

| | Baseline, n (%) | | |
|---|---|---|---|
| **Follow-up:** | **Day work** | **Shift work** | **Total** |
| **Day work** | 706 (84%) | 122 (12%) | 828 |
| **Shift work** | 138 (16%) | 897 (88%) | 1,035 |
| **Total** | 844 (100%) | 1,019 (100%) | 1,863 |

**Table S3**. Agreement statistics of baseline and retrospectively reported shift work status using three-category shift work variable.

| | **Baseline, n (%)** | | | |
|---|---|---|---|---|
| **Follow-up report** | Day work | Occasional (2–3 times/ year) | Shift work | Total |
| Day work | 516 (80%) | 69 (35%) | 80 (8%) | 665 |
| Occasional shift work | 58 (9%) | 63 (32%) | 42 (4%) | 163 |
| Shift work | 71 (11%) | 67 (34%) | 897 (88%) | 1,035 |
| Total | 645 | 199 | 1,019 | 1,863 |
| **Sensitivity** | 80% (77–83) | 32% (25–39) | 88% (86–90) | |
| **Specificity** | 88% (86–90) | 94% (93–95) | 84% (81–86) | |
| **Agreement** | 79% (77–81) | | | |
| **Cohen's κ** | 0.63 (0.60–0.66) | | | |
| **Cohen's κ (weighted)** | 0.70 (0.67–0.73) | | | |
| **Bowker's p** | 0.07 | | | |

**Table S4.** Agreement between baseline and follow-up shift-work status reports, restricting disease cases to those diagnosed before follow-up questionnaire completion.

| Disease | Case | n | Agreement (95% CI) | Cohen's κ (95% CI) | McNemar's p | Sensitivity (95% CI) | Specificity (95% CI) |
|---|---|---|---|---|---|---|---|
| Cancer | No | 1,846 | 86% (84–88) | 0.72 (0.69–0.75) | 0.26 | 88% (86–90) | 84% (81–86) |
| | Yes | 17 | 88% (64–99) | 0.68 (0.28–1.00) | 0.16 | 86% (57–98) | 100% (29–100) |
| Cardiomet. disease | No | 1,849 | 86% (84–88) | 0.72 (0.69–0.75) | 0.32 | 88% (86–90) | 84% (81–86) |
| | Yes | 14 | 71% (42–92) | 0.43 (-0.04–0.90) | 1.00 | 71% (29–96) | 71% (29–96) |

Sensitivity analysis restricted to cases diagnosed before follow-up completion showed a similar pattern to the main analysis: no evidence of asymmetric misclassification for disease subgroups, and lower agreement among cardiometabolic cases than non-cases. Confidence intervals were wide given small subgroup sizes.
